# Differential Involvement of the Locus Coeruleus in Early- and Late-Onset Alzheimer’s Disease: A Potential Mechanism of Clinical Differences?

**DOI:** 10.1101/2020.11.01.20224139

**Authors:** Corey J. Bolton, Joyce W. Tam

## Abstract

Early-onset Alzheimer’s disease (EOAD) has been associated with an increased likelihood of atypical clinical manifestations such as attentional impairment, yet the cause of this heterogeneity remains unclear. The locus coeruleus (LC) is implicated early in Alzheimer’s disease pathology and is associated with attentional functioning. This study investigated post-mortem atrophy of the LC in EOAD and late-onset Alzheimer’s disease (LOAD) in a large, well-characterized sample. Results show nearly four times greater likelihood of higher LC atrophy in EOAD as compared to LOAD after controlling for other measures of pathological progression (*p* < .005). Follow-up analyses within the EOAD group revealed that compared to those who displayed mild or no LC atrophy at autopsy, those with moderate-severe atrophy of the LC displayed significantly worse performance on various baseline measures of attentional functioning (*p* < .05), despite similar overall cognition (*p* = .25). These findings suggest the LC is an important potential driver of clinical and pathological heterogeneity in EOAD.

## 1. Introduction

Alzheimer’s disease (AD) is increasingly recognized as a heterogenous disease with various clinical presentations and neuropathological subtypes (Murray et al., 2011). The underlying causes of these clinical and pathological differences have yet to be elucidated; however, age of disease onset may be an important element to consider. Early-onset Alzheimer’ disease (EOAD), defined as sporadic AD with onset at age 65 or younger, has been associated with increased likelihood of atypical clinical phenotypes and pathological features that suggest that this condition could be distinguished from late-onset AD (LOAD) on the basis of more than age alone (van der Flier et al., 2011). As a group, patients with EOAD have been shown to have more widespread atrophy, more impairment in non-memory cognitive domains, and more rapid cognitive decline when compared to patients with LOAD (Barnes et al., 2015; Bolton, et al., 2018; Bolton & Pyykkonen, 2019; Dickerson et al., 2016; Joubert et al., 2016; Koedam et al., 2010; Palasí et al., 2015; Smits et al., 2015). In a recent retrospective review, it was reported that up to two-thirds of patients with EOAD present early in their disease course with a non-amnestic cognitive impairment, compared to approximately one-eighth of patients with LOAD (Mendez et al., 2012).

While atypical features in EOAD have been readily described in the existing literature, there has been very few studies investigating the factors that may contribute to these differences in AD. Examination of the underlying pathology in EOAD would be critical for the development of effective, targeted treatments. Without a clear understanding of the factors that influence the clinical heterogeneity in EOAD, there may be an increased likelihood of misdiagnosis that could prevent or delay these patients from receiving appropriate treatment or enrolling in clinical trials. Further, misdiagnosis or delay to accurately diagnose individuals with EOAD could contribute to inaccurate prognosis, which has significant ramifications for patients as well as their families and caregivers.

Various anatomical structures have been implicated in EOAD (e.g., greater atrophy of the precuneus and posterior cingulate; Dickerson et al., 2016; Frisoni et al., 2007; Karas et al., 2007; Möller et al., 2013; Verclytte et al., 2016). To our knowledge, no studies to date have examined the role of locus coeruleus (LC) pathology in EOAD despite evidence showing that the LC is the earliest site of pathological tau aggregation in AD (Braak et al., 2011). The LC is a small, blue colored nucleus in the brainstem that is the primary site for norepinephrine producing neurons. It has been demonstrated that the locus coeruleus-norepinephrine (LC-NE) system is essential for cognitive functions including attention and awareness (Aston-Jones & Cohen, 2005; Coull et al., 1999; Murphy et al., 2011; Sara, 2009) as well as emotional functioning (Moret & Briley, 2011). EOAD has been associated with attentional impairment (Bolton et al., 2018; D. Jacobs et al., 1994; Palasí et al., 2015; Smits et al., 2015) and up to 85% of patients with EOAD display neuropsychiatric symptoms (Ballarini et al., 2016); thus, the clinical presentation of EOAD suggests potential dysfunction of the LC-NE system. The present study investigated the role that the LC may play in the clinical heterogeneity seen in individuals with AD pathology. More specifically, we hypothesized that a greater level of atrophy in the LC at autopsy was seen in patients with EOAD when compared to others with LOAD. Further, it is hypothesized that differences in antemortem cognitive functioning would be observed between patients with EOAD who exhibit greater LC atrophy at autopsy and those patients with EOAD who have less postmortem LC atrophy.

## 2. Method

### 2.1 Participants

Data in the present study was obtained through the National Alzheimer’s Coordinating Center (NACC) database. The database contains patient data from the Alzheimer’s disease centers (ADCs) across the United States. Each ADC utilizes a standardized evaluation protocol and longitudinal study design. While the overall database includes subjects with AD and other related dementias, only data from participants with autopsy-confirmed AD pathology who made their initial visit to ADCs between September 2005 and November 2018 were included in the present study (N = 787). Inclusion criteria were: (1) autopsy evidence of AD pathology defined as neurofibrillary tangles consistent with Braak stages 3 through 6, as well as moderate to frequent neuritic plaques; and (2) neuropathological examination of locus coeruleus hypopigmentation. Participants were assigned to one of two groups for the main analysis based on their age of clinician-determined symptom onset: (1) EOAD (age ≤ 65; N = 115); and (2) LOAD (age > 65; N= 672). Participants from the EOAD group were further classified in one of two groups based on their degree of locus coeruleus hypopigmentation previously established in the database: (1) less LC atrophy (i.e., none to mild hypopigmentation); and (2) greater LC atrophy (i.e., moderate to severe hypopigmentation). Clinical data was obtained during participants’ initial visit at an ADC and time to death was measured as the duration between this initial visit and death.

### 2.2 Measures

#### 2.2.1 Neuropathology

Neuropathology data was reported using standard NACC neuropathology reporting forms and included pathologist report of Braak staging and frequency of neuritic plaques. To determine levels of atrophy of the locus coeruleus, neuropathologists’ rating of hypopigmentation was used; this has been shown to be a marker for degeneration of neuromelanin containing neurons and serves as a proxy for atrophy in this region (Ellison et al., 2013).

#### 2.2.2 Neuropsychological Tests

The Uniform Data Set (UDS) is a systematic and centralized method of assessing patients seen across ADCs since 2005. The present study utilized the neuropsychological test battery from the UDS. The neuropsychological test battery of the UDS consists of brief measures of attention, processing speed, executive function, episodic memory, and language (Weintraub et al., 2009). All neuropsychological measures were initially reported as raw scores and were subsequently converted to age-, sex-, and education-corrected z-scores using a regression based norms calculator provided by NACC (Shirk et al., 2011).

The Mini-Mental State Examination (MMSE; Folstein et al., 1975) was used to measure global cognitive functioning. To measure attention, the digit span forward and digit span backward subtests of the Wechsler Memory Scale-Revised (WMS-R; Wechsler, 1987b) were utilized. Also from the WMS-R, the Logical Memory Story A, immediate and delayed recall trials were used as a measure of episodic memory. Processing speed was assessed using the digit symbol subtest from the Wechsler Adult Intelligence Scale-Revised (WAIS-R; Wechsler, 1987a), as well as part A of the Trail Making Test (Reitan, 1992). Part B of the Trail Making Test was included as a measure of executive functioning. To assess language abilities, an abbreviated, 30-item version of the Boston Naming Test (Kaplan et al., 1983) was used along with animal and vegetable list generation tasks.

### 2.3 Statistical Analyses

Data were analyzed using IBM SPSS version 25. Independent t-tests or chi-square tests were used for between group comparisons. To investigate the influence of age group on degree of locus coeruleus hypopigmentation, ordinal logistic regression with proportional odds was used. Pearson correlational analysis was used to determine potential covariates to include in the ordinal logistic regression model.

## 3. Results

Participant characteristics were reported in Table 1. More specifically, individuals with EOAD were significantly younger at the initial visit when compared to those with LOAD. In addition, at the time of autopsy, the EOAD group was more likely to be APOE ε4 positive, had a higher average Braak staging score, had more frequent neuritic plaques, and had more severe hypopigmentation of the locus coeruleus. To determine which covariates to include in the regression model, correlational analysis between hypopigmentation of the locus coeruleus and potentially influential variables was conducted. Braak stage, frequency of neuritic plaques, number of ε4 alleles, and time to death were all significantly correlated with hypopigmentation of the locus coeruleus (see Table 2).

**Table 1.**
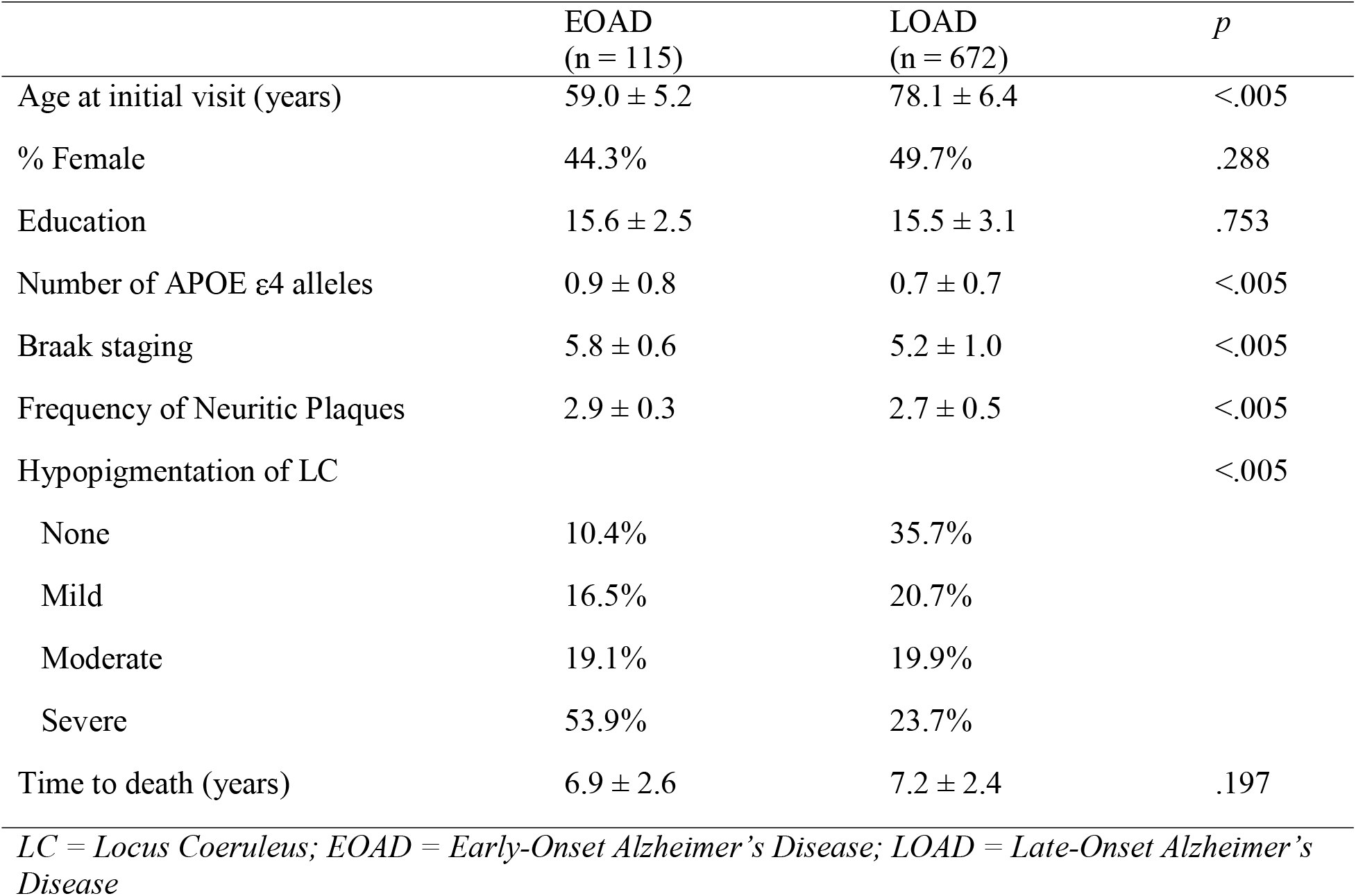
Sample Characteristics

|  | EOAD<br>(n = 115) | LOAD<br>(n = 672) | *p* |
| --- | --- | --- | --- |
| Age at initial visit (years) | 59.0 ± 5.2 | 78.1 ± 6.4 | <.005 |
| % Female | 44.3% | 49.7% | .288 |
| Education | 15.6 ± 2.5 | 15.5 ± 3.1 | .753 |
| Number of APOE ε4 alleles | 0.9 ± 0.8 | 0.7 ± 0.7 | <.005 |
| Braak staging | 5.8 ± 0.6 | 5.2 ± 1.0 | <.005 |
| Frequency of Neuritic Plaques | 2.9 ± 0.3 | 2.7 ± 0.5 | <.005 |
| Hypopigmentation of LC |  |  | <.005 |
| None | 10.4% | 35.7% |  |
| Mild | 16.5% | 20.7% |  |
| Moderate | 19.1% | 19.9% |  |
| Severe | 53.9% | 23.7% |  |
| Time to death (years) | 6.9 ± 2.6 | 7.2 ± 2.4 | .197 |

*LC = Locus Coeruleus; EOAD = Early-Onset Alzheimer's Disease; LOAD = Late-Onset Alzheimer's Disease*

**Table 2.**
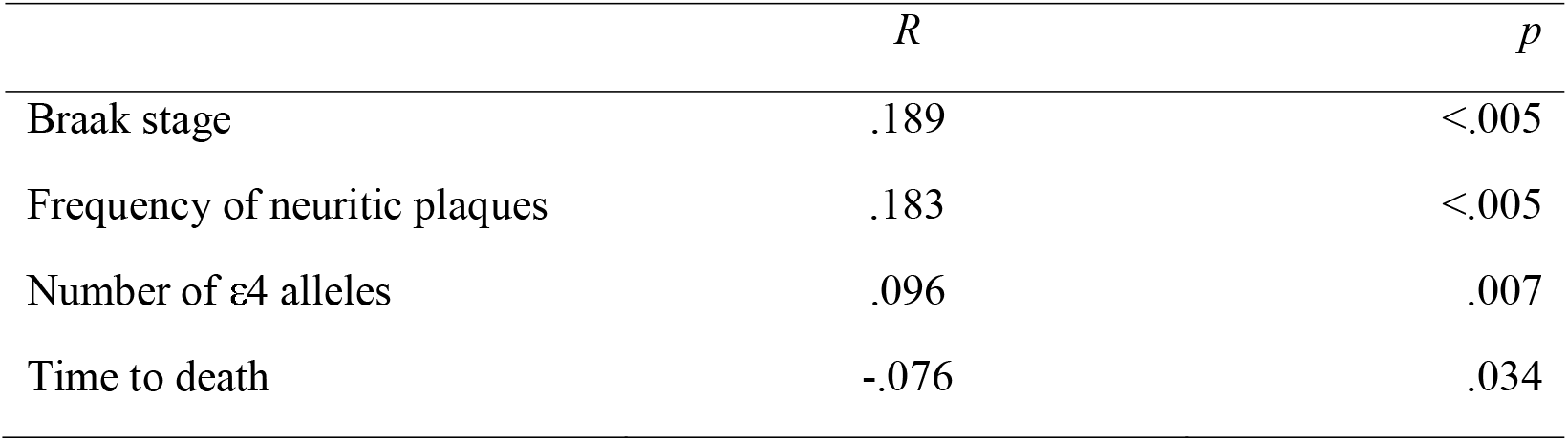
Correlations of potential covariates with hypopigmentation of locus coeruleus

|  | *R* | *p* |
| --- | --- | --- |
| Braak stage | .189 | <.005 |
| Frequency of neuritic plaques | .183 | <.005 |
| Number of ε4 alleles | .096 | .007 |
| Time to death | -.076 | .034 |

As shown in Table 3, an ordinal logistic regression with proportional odds was run to determine the effect of age at symptom onset on hypopigmentation of the locus coeruleus at autopsy after controlling for time to death, APOE genotype, and pathological markers of tau and amyloid burden. There were proportional odds, as assessed by a full likelihood ratio test comparing the fitted model to a model with varying location parameters, χ2(10) = 18,23 *p* > .05. The deviance goodness-of-fit test indicated that the model was a good fit to the observed data, χ2(691) = 723.84, *p* > .05. The final model statistically significantly predicted the dependent variable over and above the intercept-only model, χ2(5) = 81.05, *p* < .001. Age group was the best predictor of hypopigmentation of the LC in comparison to the other covariates (regression coefficient = 1.37). The odds of patients in the EOAD group having higher hypopigmentation of the LC was 3.94 (95% CI, 2.60 to 5.96) times that for patients in the LOAD group, χ2(1) = 42.02, *p* <.005.

**Table 3.** Results of ordinal logistic regression

|  | Regression coefficient | Standard error | Wald | *P* | OR | 95% CI |
| --- | --- | --- | --- | --- | --- | --- |
| Braak stage | .16 | .09 | 3.30 | .069 | 1.17 | 0.99, 1.40 |
| Frequency of neuritic plaques | .46 | .17 | 7.19 | .007 | 1.58 | 1.13, 2.21 |
| Time to death | -.04 | .03 | 1.53 | .217 | 0.96 | 0.91, 1.02 |
| Number of ε4 alleles | -.02 | .12 | 0.02 | .879 | 0.98 | 0.78, 1.24 |
| Age group = EOAD | 1.37 | .21 | 42.02 | <.005 | 3.94 | 2.60, 5.96 |

*OR = Odds Ratio; CI = Confidence Interval***Table 4***Characteristics of EOAD group*

To further understand the effects of LC integrity on cognition, participants in the EOAD group were divided into two groups based on degree of LC atrophy. Characteristics of these two groups are presented in Table 4. No group differences were observed on any demographic variables, APOE genotype, time to death, or measures of tau and amyloid pathological burden.

**Table 4.** Characteristics of EOAD group

|  | Less LC Atrophy (n = 31) | Greater LC Atrophy (n = 84) | *p* |
| --- | --- | --- | --- |
| Age at initial visit (years) | 59.2 ± 5.9 | 58.9 ± 5.0 | .769 |
| % Female | 35.5% | 47.6% | .245 |
| Education | 15.0 ± 2.6 | 15.8 ± 2.5 | .089 |
| Number of APOE ε4 alleles | 0.9 ± 0.8 | 0.9 ± 0.8 | .692 |
| Braak staging | 5.7 ± 0.7 | 5.8 ± 0.5 | .605 |
| Frequency of Neuritic Plaques | 2.8 ± 0.4 | 2.9 ± 0.3 | .155 |
| Time to death (years) | 7.2 ± 2.3 | 6.8 ± 2.7 | .461 |

*LC = Locus Coeruleus*

Comparisons between the greater and less LC atrophy groups on individual neuropsychological measures is presented in Table 5. The two groups did not differ on a measure of global cognition (i.e., MMSE). However, those with greater locus coeruleus atrophy performed significantly worse on WMS-R Logical Memory I, Digit Span Forward, Digit Span Backward, and one trial of semantic fluency (i.e., Animals) with effect sizes on these measures ranging from .39 to .58. The two groups did not significantly differ on any other measure, including the delayed recall trial of Logical Memory.

**Table 5.** Comparison of neuropsychological test performance in EOAD group

|  | Less LC Atrophy<br>(n = 31) | Greater LC Atrophy<br>(n = 84) | *p* | Cohen's *d* |
| --- | --- | --- | --- | --- |
| Logical Memory I | -1.43 ± 1.51 | -1.93 ± 1.02 | .042 | 0.39 |
| Logical Memory II | -1.52 ± 1.46 | -1.91 ± 0.96 | .107 | 0.33 |
| Digit Span Forward | -0.67 ± 1.11 | -1.20 ± 1.25 | .039 | 0.45 |
| Digit Span Backward | -0.98 ± 1.02 | -1.40 ± 0.90 | .032 | 0.44 |
| Animals | -1.20 ± 0.97 | -1.80 ± 1.09 | .008 | 0.58 |
| Vegetables | -1.18 ± 1.18 | -1.25 ± 1.12 | .794 | 0.05 |
| Trail Making Test Part A | -2.29 ± 3.24 | -3.15 ± 3.06 | .189 | 0.27 |
| Trail Making Test Part B | -2.55 ± 2.32 | -3.22 ± 2.05 | .166 | 0.31 |
| Coding | -2.24 ± 1.59 | -2.51 ± 1.57 | .416 | 0.17 |
| Boston Naming Test | -1.17 ± 1.55 | -1.50 ± 2.26 | .463 | 0.16 |
| MMSE | 24.26 ± 4.46 | 23.19 ± 4.35 | .248 | 0.24 |

*Note: Neuropsychological test scores other than MMSE are age-, sex-, and education-corrected z-scores; raw scores are presented for the MMSE. EOAD = Early-Onset Alzheimer's Disease; LC = Locus Coeruleus; MMSE = Mini-Mental State Examination*

## 4. Discussion

The present study investigated differences in locus coeruleus atrophy at autopsy between patients with early- and late-onset Alzheimer’s disease using the NACC dataset. Patients in the early-onset group were nearly four times as likely to have higher atrophy of the locus coeruleus than those in the late-onset group. These results remained significant after controlling for time between initial clinic visit and death, APOE genotype, and other markers of pathological severity. A secondary analysis comparing baseline neuropsychological test performance between EOAD patients with less LC atrophy and those with greater LC atrophy at autopsy revealed significant differences on measures of verbal immediate memory, auditory attention span, working memory, and verbal fluency with the greater LC atrophy group performing worse on each of these measures despite similar levels of global cognition. These findings suggest that patients with EOAD are more likely to have greater involvement of the locus coeruleus, which may account for some of the differences in clinical manifestation of AD pathology seen between EOAD and LOAD.

It has been shown that patients with EOAD display greater deficits in non-memory cognitive domains than those patients with LOAD (Barnes et al., 2015; Bolton, et al., 2018; Bolton & Pyykkonen, 2019; Dickerson et al., 2016; Joubert et al., 2016; Koedam et al., 2010; Palasí et al., 2015; Smits et al., 2015). Despite the well-established clinical differences, there remains little understanding as to the cause of these differences. The current study extended the existing literature to show that individuals with EOAD have greater involvement of the LC than those with LOAD. Impairment in attention has previously been suggested to be one of the key features that distinguish between EOAD and LOAD (Bolton et al., 2018; Jacobs et al., 1994; Palasí et al., 2015). Consistently, we found that among individuals with EOAD, those with more LC atrophy at autopsy performed more poorly on baseline cognitive tasks involving heavy attentional demands than those with less LC atrophy. Our finding suggests that the severity of LC atrophy could play a role in the clinical phenotype observed among individuals with EOAD. This finding is consistent with past studies that showed the LC-NE system as vital for attention and awareness (Aston-Jones & Cohen, 2005; Coull et al., 1999; Murphy et al., 2011; Sara, 2009). Thus, the present study identifies a potential mechanism for these clinical differences and provides a testable hypothesis for future investigations.

In accordance with unique cognitive features, past research has identified distinct patterns of atrophy in EOAD when compared to LOAD. Specifically, it has been noted that EOAD is typically associated with more widespread atrophy and decreased functional connectivity that involves numerous brain regions (Adriaanse et al., 2014; Chung et al., 2016; Frisoni et al., 2007; Gour et al., 2014). Recent work has investigated the role of the LC in the pathogenesis of AD and has suggested that neurofibrillary tangles first appear in the LC and that this region may serve as a seed for further propagation of pathology (Braak et al., 2011; Satoh & Iijima, 2019). The LC has widespread efferent projections throughout the cortex; thus, greater involvement of the LC in EOAD may explain the more widespread pathology often seen in these patients. In our sample, Braak stage and frequency of neuritic plaques did not differ between those with lesser or greater LC atrophy; both groups showed end-stage AD pathological spread on average at autopsy. Future studies using in-vivo imaging methods would be better suited to determine the validity of this hypothesis.

In addition to a unique pattern of cognitive dysfunction and regional brain atrophy, EOAD has been associated with a faster rate of decline (Jacobs et al., 1994; Vlies et al., 2009). This feature of EOAD could also be associated with greater involvement of the LC. Wilson and colleagues (2013) found that patients who had lower density of noradrenergic neurons in the LC at autopsy showed a greater rate of antemortem cognitive decline. The authors suggested that integrity of the LC-NE system could serve as a biological mechanism of reserve. The greater atrophy seen in the LC of EOAD patients in this study could reflect this decreased neural reserve and thereby provide a potential explanation for the increased rate of cognitive decline in EOAD.

It should be noted that a significant proportion of patients with EOAD did not show high levels of LC atrophy upon autopsy, despite similar demographic backgrounds and time to death. There has been a burgeoning interest in identifying subtypes of AD, with this work leading to the identification of at least three pathologically defined subtypes of AD including limbic-predominant, hippocampal-sparing, and typical AD (Murray et al., 2011). While this study did not examine regional density of neurofibrillary tangles and atrophy, differing patterns of LC atrophy in patients with EOAD in this group could represent different “subtypes” of AD. This could explain the differences seen in the clinical presentation of EOAD, with a sizable proportion of these patients displaying a typical, amnestic cognitive profile.

These findings have a number of possible implications for the field. First, a nuanced understanding of the different clinical and pathological presentations of AD would allow for early and accurate diagnosis. Patients who are under the age of 65 and are experiencing progressive cognitive decline may often be misdiagnosed, especially if they do not have a typical, amnestic cognitive impairment. Early and accurate diagnosis will also provide greater precision in clinical trial selection and possible treatments.

Next, these findings suggest, in accord with past research, that the LC is an important region for cognitive functioning in patients with AD, and thus an area of consideration for pharmacological intervention. While most studies to date have focused on norepinephrine therapy as a treatment for neuropsychiatric symptoms in late dementia, there is recent increased interest in repurposing the many available drugs that target the LC-NE system for use in early-phase AD clinical trials (Chalermpalanupap et al., 2017). Atomoxetine, a norepinephrine reuptake inhibitor, has been shown to improve attention, memory, and executive functions in nonhuman primate studies (Callahan et al., 2019) and human studies of adults with ADHD (Ni et al., 2013), but did not improve cognition as an augment to cholinesterase-inhibitor therapy in patients with mild-to-moderate AD (Mohs et al., 2009). However, as in many AD clinical trials to date, it is possible that treatment was initiated too late and the effect of such medications at early disease stages is yet to be seen. Our findings suggest that patients who go on to have significant LC atrophy post-mortem display cognitive deficits consistent with LC-NE system dysfunction at their initial visit to a memory clinic, thus highlighting the potential early effects of LC involvement in AD, and EOAD in particular. Initiating treatment to promote norepinephrine activity at the preclinical stages may stall or halt progression of cognitive decline. Recent work has highlighted the LC-NE system’s anti-inflammatory functions (Chalermpalanupap et al., 2017; Feinstein et al., 2016; H. I. L. Jacobs et al., 2019) and its role in neurogenesis (Chalermpalanupap et al., 2013; Leanza et al., 2018). Promoting activity of the LC-NE system early in the disease course could potentially interrupt some of the key drivers of AD progression.

While this study has a number of strengths including being the first of its kind to investigate LC involvement and its potential cognitive effect in EOAD, careful consideration of potential confounding variables, and important clinical implications, it is not without its weaknesses. The method of determining LC atrophy was qualitative in nature, being based on gross examination by trained pathologists and does not provide the same level of precision as unbiased stereological methods. Further, there was no in-vivo determination of LC involvement; thus, no causal statements regarding the role of LC involvement in cognitive functioning can be made in this sample. Despite these limitations, this study represents an important first step for understanding the role of the LC in EOAD.

Future research should consider investigating the pathological burden of the LC in patients with EOAD in-vivo using neuroimaging methods. These patients should undergo serial assessment using neuroimaging and measures of cognitive functioning to understand if there exist different disease trajectories based on involvement of the LC. In addition, these findings suggest that further investigation of existing noradrenergic medications may be of benefit in patients with EOAD and may warrant clinical investigation.

## Data Availability

All data are publicly available through the National Alzheimer's Coordinating Center.

https://www.alz.washington.edu

## Acknowledgements

The NACC database is funded by NIA/NIH Grant U01 AG016976. NACC data are contributed by the NIA-funded ADCs: P30 AG019610 (PI Eric Reiman, MD), P30 AG013846 (PI Neil Kowall, MD), P30 AG062428-01 (PI James Leverenz, MD) P50 AG008702 (PI Scott Small, MD), P50 AG025688 (PI Allan Levey, MD, PhD), P50 AG047266 (PI Todd Golde, MD, PhD), P30 AG010133 (PI Andrew Saykin, PsyD), P50 AG005146 (PI Marilyn Albert, PhD), P30 AG062421-01 (PI Bradley Hyman, MD, PhD), P30 AG062422-01 (PI Ronald Petersen, MD, PhD), P50 AG005138 (PI Mary Sano, PhD), P30 AG008051 (PI Thomas Wisniewski, MD), P30 AG013854 (PI Robert Vassar, PhD), P30 AG008017 (PI Jeffrey Kaye, MD), P30 AG010161 (PI David Bennett, MD), P50 AG047366 (PI Victor Henderson, MD, MS), P30 AG010129 (PI Charles DeCarli, MD), P50 AG016573 (PI Frank LaFerla, PhD), P30 AG062429-01(PI James Brewer, MD, PhD), P50 AG023501 (PI Bruce Miller, MD), P30 AG035982 (PI Russell Swerdlow, MD), P30 AG028383 (PI Linda Van Eldik, PhD), P30 AG053760 (PI Henry Paulson, MD, PhD), P30 AG010124 (PI John Trojanowski, MD, PhD), P50 AG005133 (PI Oscar Lopez, MD), P50 AG005142 (PI Helena Chui, MD), P30 AG012300 (PI Roger Rosenberg, MD), P30 AG049638 (PI Suzanne Craft, PhD), P50 AG005136 (PI Thomas Grabowski, MD), P30 AG062715-01 (PI Sanjay Asthana, MD, FRCP), P50 AG005681 (PI John Morris, MD), P50 AG047270 (PI Stephen Strittmatter, MD, PhD).

## Notes

### Competing Interest Statement

The authors have declared no competing interest.

### Funding Statement

No external funding was received in support of this project.

### Author Declarations

As determined by the University of Washington Human Subjects Division, the NACC database itself is exempt from IRB review and approval because it does not involve human subjects, as defined by federal and state regulations. However, all contributing ADCs are required to obtain informed consent from their participants and maintain their own separate IRB review and approval from their institution prior to submitting data to NACC.

